# Optimisation of steatotic liver disease screening algorithm for resource-poor settings using machine learning

**DOI:** 10.64898/2026.06.09.26355306

**Authors:** Chamila Mettananda, Kaveesha Sivasumithran, Ranaweera L Lakmali, Anjalika Madhubhashini, Chamila Ranawaka, Arunasalam Pathmeswaran, Anuradha Dassanayake

**Affiliations:** Faculty of Medicine, University of Kelaniya, Ragama, Sri Lanka; Faculty of Science, University of Colombo, Colombo 03, Sri Lanka; North Colombo Teaching Hospital, Ragama, Sri Lanka

**Author notes:** Corresponding author, (CM). These authors contributed equally to this work (Chamila Mettananda, Kaveesha Sivasumithran).

**Keywords:** SLD, steatotic liver disease, MASLD, significant fibrosis, prediction, machine learning, VCTE, ESAL

## Abstract

**Background:** The European Association for the Study of the Liver (ESAL) - Steatotic Liver Disease (SLD) screening algorithm involves two steps; initial screening with FIB-4 followed by referral for vibration-controlled transient elastography (VCTE) in patients likely to have significant fibrosis (SF). However, VCTE is not widely available in resource-limited settings.

**Aim:** To optimise the EASL SLD screening algorithm for resource-poor settings using machine learning (ML).

**Methods:** We analysed data from 964 adults aged ≥35 years who underwent VCTE at a tertiary referral centre in Sri Lanka between November 2024 and 2025. Multiple ML models using different methods and variable combinations were trained on 80% of the dataset and tested on the remaining 20%. Best models were selected based on performance and externally validated using data from 430 patients who underwent VCTE before November 2024. Model performance was compared with the FIB-4 using confusion matrices.

**Results:** A Random Forest model incorporating age, AST, ALT, and platelet count separately, rather than using FIB-4, outperformed. The all-variable ML model showed the best predictive performance for SF, with accuracy of 77.2%, recall of 0.762, precision of 0.778, and AUC-ROC of 0.818. The variables used in the model, in descending order of feature importance, were AST, platelet count, BMI, ALT, age, diabetes mellitus, hypertension, dyslipidaemia, sex, family history, hypothyroidism, diabetes complication and smoking.

External validation demonstrated 75.1% accuracy and an AUC of 0.779. When used as the first step of the SLD screening algorithm, the all-variable ML model identified 37 (17.1%) additional true positives and reduced false-negative diagnoses by 50% compared with FIB-4.

**Conclusions:** ML-based models were more effective than the FIB-4 score as the first-line screening tool for VCTE referral, substantially improving the identification of patients with significant fibrosis in this South Asian cohort.

## Introduction

Steatotic liver disease (SLD) is the most common chronic liver disease in the world, affecting 30-35% [1–4]. It is a spectrum of disease spanning from simple steatosis to cirrhosis [5–7]. Around 10-20% of patients with SLD progress to advanced chronic liver cell disease (ACLD) over 15-20 years [3, 6–9]. The stage of liver fibrosis predicts disease progression. Patients with significant liver fibrosis (SF) are likely to progress to advanced fibrosis (AF) [10–12]. Therefore, early detection and initiation of liver-directed therapy at the SF stage is recommended [13–16].

The European Society for the Study of Liver (EASL) guideline recommends annual screening of patients with diabetes or SLD using a two-step process. First, using FIB-4score, a non-invasive biochemical test to identify those likely to have SF, and subsequently imaging those likely to have SF using vibration-controlled transient elastography (VCTE) or Magnetic resonance elastography (MRE) as the second step [14, 17–23].

However, neither VCTE nor MRE is readily available in resource-limited settings; therefore, it is helpful to improve selection for VCTE further. Current recommendation in European guidelines is to refer patients with FIB-4score ≥ 1.3 (≥2 in those above 65 years old) for VCTE, but FIB-4score has not been validated in most South Asians [15]. Furthermore, machine learning (ML) models have been shown to perform better at predicting SF than other non-invasive tests such as FIB-4, FAST, and NFS across European, American, Chinese, and some Asian populations [24–34].

Therefore, we aimed to develop ML-based models to predict SF using medical history, anthropometric measurements, and freely available blood investigations, and to compare the new models’ predictions with those of the FIB-4 score as the first step in the screening algorithm for SLD to optimise the utilisation of limited VCTE facilities in resource-limited settings.

## Methods

We used de-identified FibroScan Registry data from patients with VCTE at a tertiary care referral centre in Sri Lanka from 1^st^ November 2024 to 1^st^ November 2025. This is the only state-sector centre in Sri Lanka with VCTE. We extracted data on 1^st^ November 2025 from patients aged 35 years or older with ultrasonographic (US) evidence of SLD and complete data to calculate FIB-4, including serum aspartate aminotransferase (AST), alanine aminotransferase (ALT), and platelet count. We excluded patients with US evidence of cirrhosis, and those with SLD due to causes other than metabolic-associated steatotic liver disease (MASLD), such as methotrexate, tamoxifen, and unsafe alcohol intake or hepatitis. We calculated the FIB-4 score of the patients using the following equation [35].

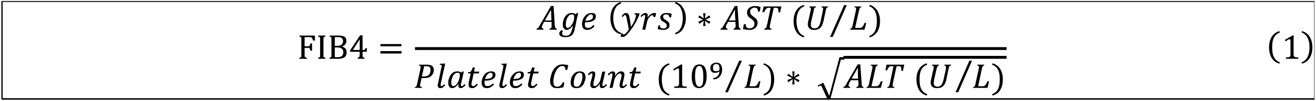

### Model development

We developed several ML models to predict SF using registry data. First, the dataset was pre-processed, including Multiple Imputation by Chained Equations (MICE) for handling missing values and label encoding to obtain a clean dataset for model building. Graphical exploratory data analysis was performed to obtain more information about the data’s trends and patterns. Different variable combinations were tested to predict SF based on variable importance and domain knowledge of SF. A slight imbalance of the response variable was noted, with 58.51% patients having SF and 41.49% not having SF. To address this, the Synthetic Minority Over-sampling Technique (SMOTE) was used, and models trained on balanced data were compared with those trained on the original data to identify the best model. Models were trained using different ML algorithms, namely, decision tree, random forest, XGBoost (eXtreme gradient boosting), k-Nearest Neighbours, Support Vector Machine, AdaBoost (adaptive boosting), and Gradient Boosting. Hyperparameter tuning was done using grid search for each algorithm. Models were built using the publicly available Google Colab ML platform and the Scikit-learn library in Python [9]. Participant data were split into two groups: training sample for model development (80%) and testing sample for internal validation (20%). The training sample was used to build the ML models, and the testing sample was used to assess the efficacy of the algorithms trained on it. The models were compared for predictive performance using accuracy, precision, and recall, and the best model was selected.

We compared the performance of the ML-based model vs the FIB-4 score in predicting SF as the first step in the screening algorithm for SLD, using the area under the receiver operating characteristic curves (AUC-ROC). Using confusion matrices, we compared the added benefit of the new ML model over the current recommendation for using the FIB-4 score.

### External validation

Next, we externally validated the new ML-based model using a separate, de-identified database of patients with MASLD who had VCTE performed before 1^st^ November 2024. We compared the performance of the new model vs the FIB-4 score in predicting SF as the first step in the screening algorithm for SLD on the external validation cohort using AUC and confusion matrices.

All experiments were conducted in accordance with the Declaration of Helsinki. Ethical clearance was granted for the FibroScan Registry (P/143/10/2023) and the external validation cohort (P/66/07/2021) by the Ethics Review Committee of the Faculty of Medicine, University of Kelaniya, Sri Lanka. Written informed consent was obtained from all the participants. The authors did not have access to information that could identify individual participants during or after data collection for this secondary analysis.

## Results

ML-based models were developed using the Fatty Liver database. The selected sample consisted of 964 adults: 44% males, mean age of 57.9 years, with 58.5% having SF confirmed on VCTE. Baseline characteristics of the study population are shown in Table 1.

**Table 1.** Baseline characteristics of the cohort used in ML-based model development and validation.

|  | Total |
| --- | --- |
|  | N = 964 |
| Age, mean (SD<sup>a</sup>), years | 57.3 (9.6) |
| Age ≤ 65 years, N<sup>b</sup> (%) | 774 (80.3) |
| Age > 65 years, N<sup>b</sup> (%) | 190 (19.7) |
| Male sex, N<sup>b</sup> (%) | 422 (43.8) |
| Hypertension, N<sup>b</sup> (%) | 396 (41.1) |
| Dyslipidaemia, N<sup>b</sup> (%) | 486 (50.4) |
| Diabetes N<sup>b</sup> (%) | 439 (45.5) |
| Diabetic complications present, N<sup>b</sup> (%) | 38 (3.9) |
| Hypothyroidism N<sup>b</sup> (%) | 82 (8.5) |
| Current smokers, N<sup>b</sup> (%) | 23 (2.4) |
| Family history of liver disease, N<sup>b</sup> (%) | 88 (9.1) |
| BMI<sup>c</sup> Mean (SD<sup>a</sup>) Kg/m<sup>2</sup> | 25.52 (4.2) |
| Overweight/ obesity (BMI<sup>c</sup> ≥ 23 Kg/m<sup>2</sup>), N<sup>b</sup> (%) | 696 (72.1) |

SD<sup>a</sup> standard deviation, N<sup>b</sup> number, BMI<sup>c</sup> Body Mass Index

Two variable combinations were used for model development: Model 1 (age, platelet count, AST, and ALT values used separately) and Model 2 (age, platelet count, AST, and ALT values replaced with the FIB-4 score). Multiple ML algorithms were trialled for each variable combination, and the performance metrics of the best-performing model for each are shown in Table 2. Model 1, developed using the Random Forest method (with unbalanced original data), had the highest performance.

**Table 2.** Comparison of machine learning models with different sets of features.

| ML model | Optimal model | Accuracy | Recall | Precision | AUC-ROC |
| --- | --- | --- | --- | --- | --- |
| Model -1 | Random Forest on unbalanced original data | 77.2% | 0.762 | 0.778 | 0.818 |
| Model-2 | Adaboosting on data balanced using SMOTE | 73.58% | 0.738 | 0.736 | 0.796 |

SMOTE: Synthetic Minority Over-sampling Technique

AUC-ROC: Area Under the Curve – Receiver Operating Curve, ML Machine learning, SMOTE: Synthetic Minority Over-sampling Technique

Model 1- age, platelet count, AST, and ALT values used separately

Model 2 - age, platelet count, AST, and ALT values replaced with the FIB-4 score

Model-1, fitted with the Random Forest method, had 77.2% accuracy, 0.762 recall, and 0.818 AUC-ROC, and was selected as the best model. The feature importances of the predictors used in Model 1, in descending order, are shown in Fig 1. AST, platelet count, BMI, ALT, and age were the top 5 factors associated with SF.

**Fig 1.**
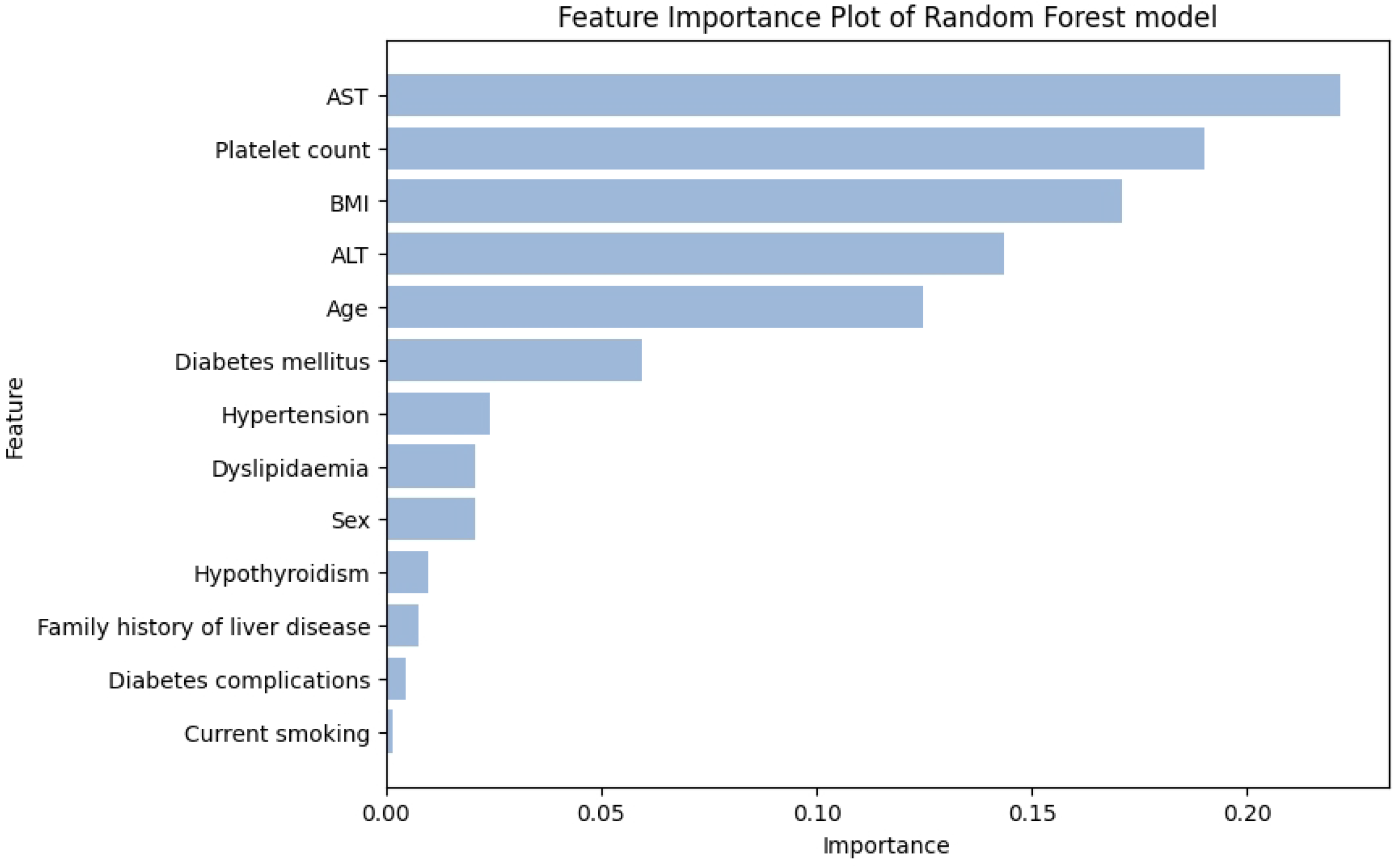
Feature importance of the variables used in ML model-1.

In search of a parsimonious model suitable for low- and middle-income countries, we built models using only five variables and 10 variables separately. The performance comparisons of the models using 5, 10, and all variables are shown in Fig 2. The model using all the variables had the highest accuracy and precision. While the 5-variable model had comparable accuracy and precision to the all-variable model, it had the highest AUC-ROC. The variables used in the 5-variable model, in descending order of feature importance, were AST, platelet count, BMI, ALT, and age.

**Fig 2.**
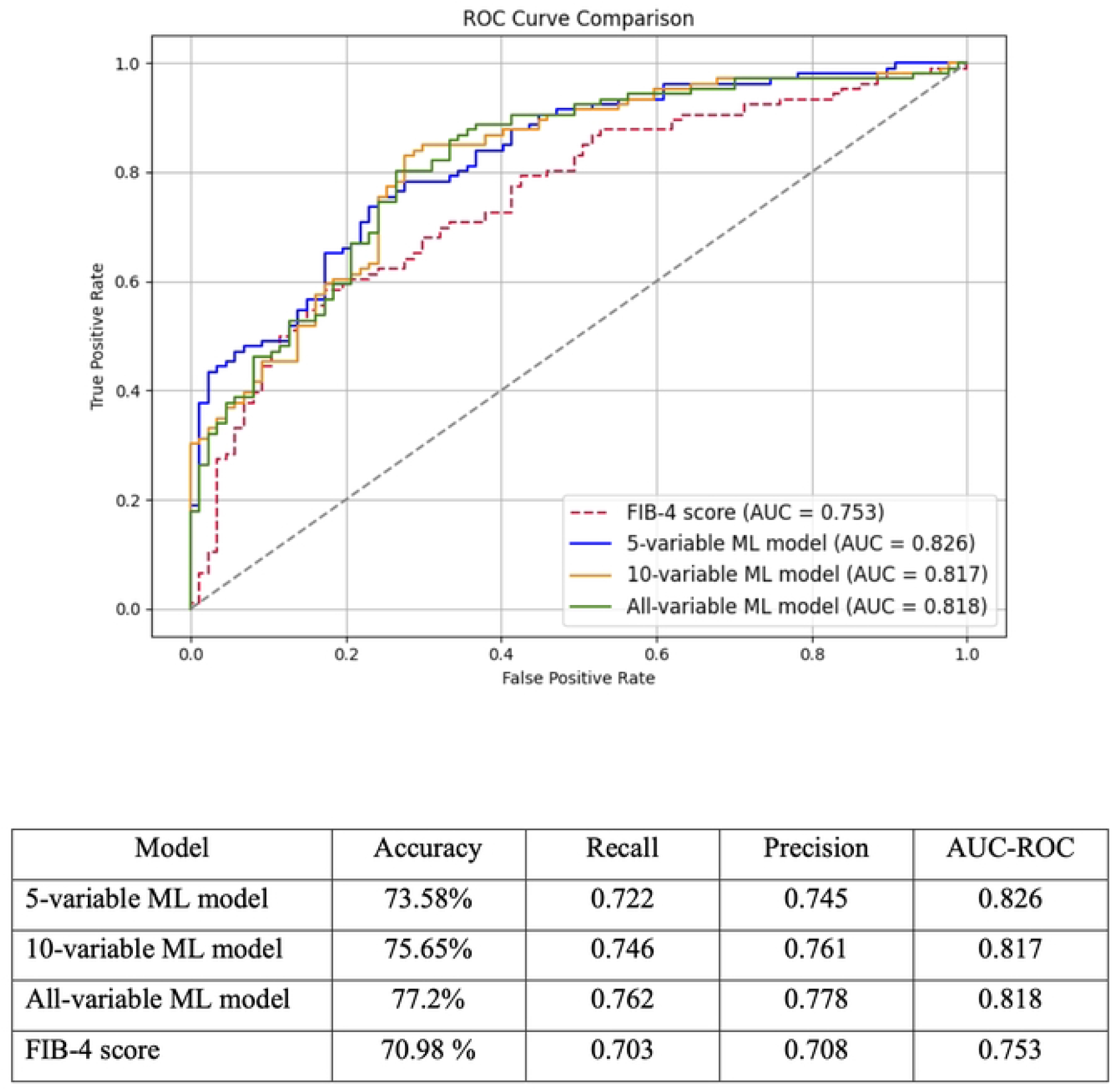
Comparison of predictive performances of the different ML models and the FIB-4 score fitted on the test sample.

AUC-ROC: Area Under the Curve – Receiver Operating Curve, ML Machine learning

### External validation

The baseline characteristics of the external validation sample are shown in Table 3.

**Table 3.** Baseline characteristics of the external validation cohort.

|  | Total |
| --- | --- |
|  | N = 430 |
| Age, mean (SD), years | 56.21 (10.9) |
| Age ≤ 65 years, N (%) | 355 (82.6) |
| Age > 65 years, N (%) | 75 (17.4) |
| Male sex, N (%) | 279 (64.88) |
| Hypertension, N (%) | 186 (43.26) |
| Dyslipidaemia, N (%) | 199 (46.28) |
| Diabetes N (%) | 220 (51.16) |
| Diabetic complications present, N (%) | 10 (2.33) |
| Hypothyroidism N (%) | 26 (6.05) |
| Family history of liver disease, N (%) | 39 (9.07) |
| Current smokers, N (%) | 52 (12.09) |
| BMI Mean (SD) Kg/m<sup>2</sup> | 25.38 (4.4) |
| Overweight/ obesity (BMI ≥ 23 Kg/m<sup>2</sup>), N (%) | 306 (71.2) |
| VCTE diagnosed significant fibrosis | 290(67.4) |

SD standard deviation, N number, BMI Body Mass Index

The performance comparisons of the 5-variable model, all-variable model, and FIB-4 score, fitted on the external validation sample, are shown in Fig 3. Both ML models, 5-variable and all-variable models, fitted on the external validation sample had better performances than the FIB-4 score in accuracy, recall, precision, and AUC-ROC. The AUC-ROC of the 5-variable model, the all-variable model, and the FIB-4 score were 0.76, 0.779, and 0.728, respectively.

**Fig 3.**
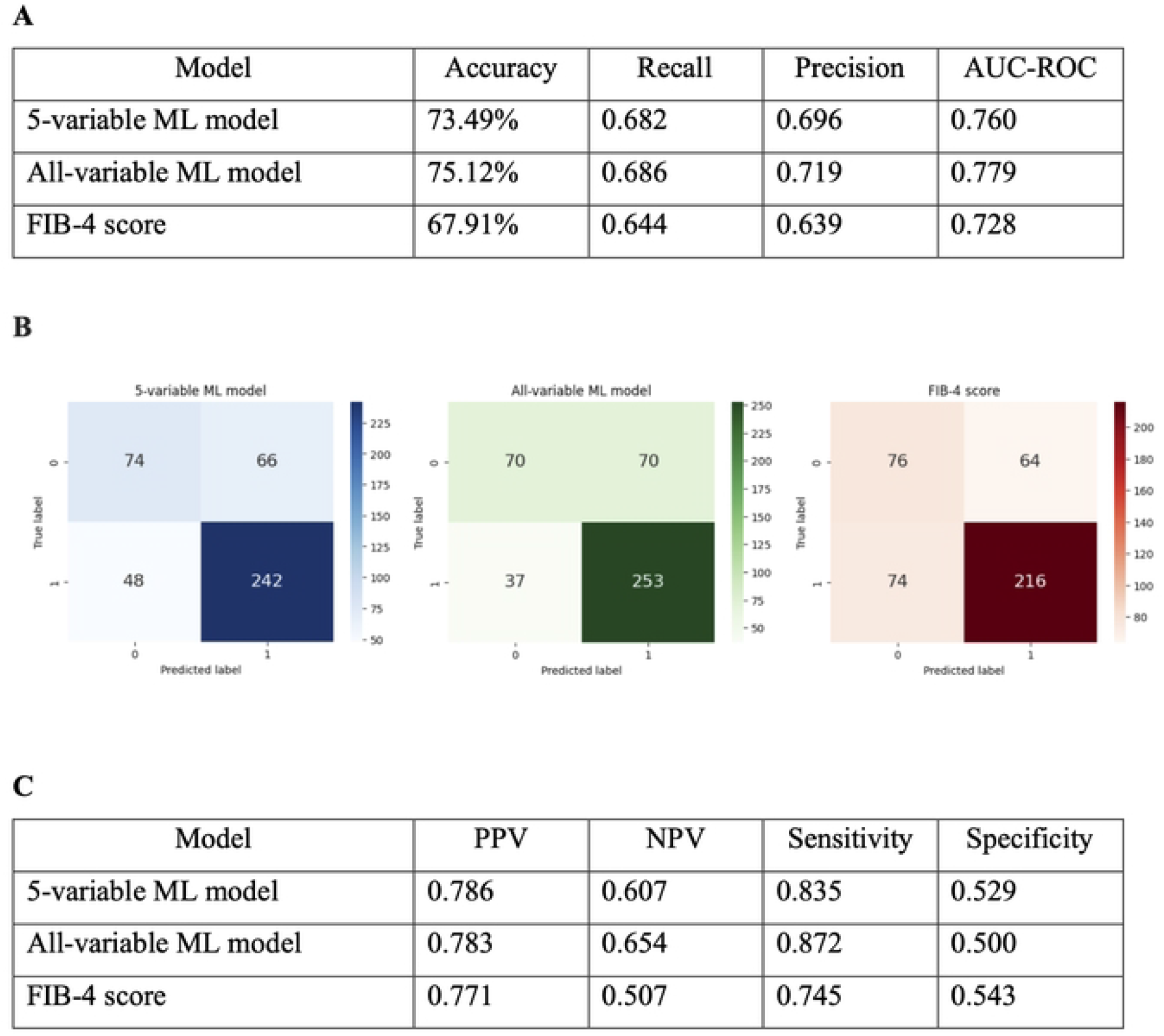
Comparison of performances of ML models and the FIB-4 score in predicting significant fibrosis in the external validation sample.

The confusion matrices for the different ML models and the FIB-4 score for predicting SF in the external validation sample, as the first step of the SLD screening algorithm, are shown in Fig 3.

Panel A – Predictive performances, Panel B – Confusion matrices, Panel C – Diagnostic accuracy measures

AUC-ROC: Area Under the Curve – Receiver Operating Curve, ML Machine learning

The all-variable ML model had a PPV of 0.783 and an NPV of 0.654, compared with 0.771 and 0.507 for the FIB-4 score (Fig 3). The ML model using all variables predicted 37 (37/216 = 17.1%) more patients with SF than the FIB-4 score. In comparison, it correctly predicted fibrosis status in 323/430 (75.1%) patients, compared with 292/430 (67.9%) with the FIB-4 score, allowing maximum utilisation of VCTE. Further, the all-variable model reduced false-negative diagnoses by 37/74 (50.0%) compared with the FIB-4 score, making it a better test to rule out SF than the FIB-4 score in this sample.

## Discussion

We observed that using models developed by machine learning as the first step of the SLD screening algorithm was more efficient than using the FIB-4 score for identifying SF in this sample of South Asians. The all-variable ML model identified 17.1% more true positives and reduced false-negative diagnoses by 50.0%. This finding is particularly valuable for resource-limited settings where VCTE is not freely available, as it allows optimal utilisation of limited resources by prioritising patients for VCTE, reducing unnecessary scans while increasing detection rates in cohorts. Even the 5-variable ML model showed higher predictive performance in the sample studied and therefore offers practical benefits for use in clinical settings.

Recent literature reports promising evidence for ML-based models outperforming well-known traditional risk prediction models [30–32, 34, 36, 37]. Our findings add to the evidence, complemented by data from South Asians. Different ML models identified different parameters to predict SF. Abdominal circumference, waist circumference, chest circumference, truncal fat, body mass index, ALT, GGT, platelet count, and age were among the most predictive variables identified by other studies, especially from Western countries, and we also identified those as predictive [25–29, 32, 33, 36]. The Gut and Obesity in Asia (GO-ASIA) study found that age, AST, ALT, fasting plasma glucose, and platelet count were the top predictors of SF across eight Asian countries [33]. In addition, we identified several additional factors as being associated. We observed that, in this South Asian sample, AST, platelet count, BMI, ALT, age, and diabetes mellitus were the top 6 factors associated with SF, in descending order. In our sample, BMI had a stronger association than age, unlike the FIB-4 score. We observed that using the four variables used in THE FIB-4 score (AST, ALT, platelet count, and age) separately, instead of using the calculated FIB-4 score as variables in the ML models, was more predictive of SF, probably explaining the differential association of the same factors in South Asians and white Caucasians. Further, we noted that the presence of diabetes mellitus was a significantly important risk factor in predicting SF in this South Asian sample, which is explained by the higher prevalence of diabetes in South Asians than in white Caucasians, while the association of SF and diabetes is already reported [38]. In addition, we identified that a family history of liver disease was a significant predictor of SF in this South Asian sample. The PNPLA3 gene has been identified as a robust genetic factor for increased liver fibrosis associated with SLD, and it is observed in South Asians predisposing those with MASLD to advanced fibrosis [39–42]. This may explain the association between family history of liver disease and SF we identified in this South Asian sample.

There are several strengths in our study. We analysed data of all consecutive patients with SLD, eliminating selection bias. VCTE was performed by the same operator using the same fibro scanner, ensuring independence and avoiding operator-related bias. We studied more than 100 variables without prejudice to identify associations with SF, then selected the best predictive variables for SF. We calculated the sample size to achieve the required effect size. Medical officers prospectively interviewed patients before VCTE; therefore, data quality is good. We validated the model’s predictions in an external cohort. There are some limitations of our study to acknowledge. We studied only a Sri Lankan cohort, and therefore, findings may not be generalisable to all South Asians, but this study paves the way for the application of the same in other South Asian countries, as we identified important risk factors related to South Asians that were not identified as so important in white Caucasians.

## Conclusion

The new ML-based models developed using non-invasive, freely available data performed better than the FIB-4 score in predicting patients with SF, as the first step in the EASL SLD screening algorithm in this South Asian sample with SLD. The new models reduced false-positive VCTEs by 50% and increased the detection rate of true positives by around 17%.

## Data Availability

The Ethics Review Committee (ERC) of the Faculty of Medicine, University of Kelaniya, has not given the approval to share participants' data. Therefore, access to the" data could only be granted for a valid request to the ERC.

